# Who Wears the Face Mask? Preventive Measures Against COVID-19 in Latin America Before Vaccination

**DOI:** 10.1101/2023.02.07.23285570

**Authors:** Elisenda Rentería, Amalia Gómez-Casillas, Pilar Zueras

## Abstract

The COVID-19 pandemic outbreak imposed the use of the sanitary mask as a protective measure to reduce the spread of the pandemic, recommended by the World Health Organization. However, the use of the face mask has been uneven and determined by individual, regional, cultural, and political factors. Based on data from the Latinobarometer, we aim to understand the profile of people who used a mask in the context of the COVID-19 pandemic in 18 countries of Latin America, between October and November 2020, right before the mass vaccination campaigns. Results show that women, older people, those with higher education, being employed and not working in temporarily jobs, retirees, students, people with a centrist political ideology, and Catholics, had a higher chance of using a face mask on a regular basis. People living in Venezuela, Chile, Costa Rica and Brazil were the most likely to use face masks. These results call attention to the need to understand social forces behind the willingness to adopt non-pharmacological preventive measures in order to make them more effective in health crisis emergencies.

## Introduction

The World Health Organization (WHO) recommended the use of face-masks as one of the main measures to prevent COVID-19 transmission (1). However, the medical discourse about preventive measures is mediated by a range of factors that affect their implementation, such as gender constructions in health care (2), risk perception related to age (3) and care attitudes linked to educational level (4). Political orientation and religious affiliation are also important explanatory aspects of how the medical/epidemiological discourses are decoded (5). In the specific context of COVID-19, the role of national governments in recognizing the magnitude of the pandemic and their willingness to implement the WHO health recommendations has also been decisive. The latter has been especially relevant in Latin America, as political instability, low levels of investment in public health systems and the idiosyncrasy of each government have defined different public interventions and prevention messages sent to the population (6).

One of the studies that have addressed the individual determinants of prevention and protection behaviors against the COVID-19 pandemic, concluded that the use of the face-mask was related to the individual perception of risk, the feeling of responsibility and solidarity, cultural and religious traditions, and the need to express their own identity (7). In relation with the adoption of protective attitudes against respiratory epidemics, a study prior to COVID-19, found that women were more inclined to apply non-pharmacological preventive measures (like the use of mask, washing hands and avoid public transport), while men were more prone to pharmacological measures (get vaccinated, use of antivirals) (8). Regarding political ideology, being of right wing ideology in the United States (US) was correlated with believing that the use of mask would not prevent the expansion of the COVID-19 virus (9), and accordingly, the US was the only country were government supporters were less likely to follow prevention health guidelines against COVID-19 in a cross-country study (5), showing the importance of the government position in individual prevention attitudes.

We did not find any work analyzing mask wearing determinants in Latin America. However, an article warned about the limitations of the region to supply enough suitable masks to the entire population and that these countries did not have previous experience in the use of masks, contrary to Asian countries (10).

The use of a face-mask during the COVID-19 pandemic has not been a trivial act and it is important to understand the individual, regional, cultural and political determinants that influence this decision to obtain lessons and design more effective preventive campaigns to manage future health crises. A transnational analysis in Latin America can allow us to understand the different levels of influence that are interconnected in the individual decision to wear a mask as a protective measure against the COVID-19 virus. This study aims to discover the profile of people who wear masks, and understand the importance of the cultural and sociopolitical context of individuals in Latin America. We carried out logistic regression models controlling for all the demographic, socioeconomic and country variables, relevant to understand the determinants of preventive behaviors.

## Data and Methods

The Latinobarometer is a representative population survey conducted in 18 Latin American countries, whose field work was between October 26 and November 17, 2020. The sample includes 20,204 people, ranging from 1,000 to 1,200 adults interviewed by country. We eliminated 127 minors, 89 people who did not answer the question about the use of masks, and 3 people who did not report their educational level.

We performed a logistic regression of the binary dependent variable created from the question about the use of mask on a regular basis to not get infected with the COVID-19 virus. The model included as independent variables the gender (male/female), age (18-25; 26-40; 41-60, 61+), educational level (basic education not completed, basic education completed, completed high school, completed higher education), current occupational status (self-employed, salaried, retired or pensioned, unemployed and student), political ideology (left, center-left, center-right, right), religion (Catholic, Evangelical, others, none) and the country of residence (Argentina, Brazil, Bolivia, Chile, Colombia, Costa Rica, Dominican Republic, Ecuador, El Salvador, Guatemala, Honduras, Mexico, Nicaragua, Panama, Paraguay, Peru, Uruguay, Venezuela).

## Results

Table 1 shows the results of the multivariate regression model, indicating women use more the mask than men (OR=1.483; p-value<0.001). Younger people are less likely to use a mask, although a greater effect is not observed among the population older than 60 years (OR= 1.562; p-value <0.001) compared to those between 41 and 60 years old (OR=1.570; p-value <0.001). Those with higher educational level, have a higher chance of wearing a mask, and differences are significant for people with higher education (OR=2.161; p-value<0.001) and secondary education (OR=1.428; p-value<0.001). Employees, temporarily unemployed people, retirees and students use the mask to a greater extent than people who are self-employed. Self-perceived social class in the bivariate analysis (results not shown here) indicates that the middle class is more likely to use masks than the upper class, but in the multivariate analysis is not significant.

**Table 1.** Logistic regression over the regular use of face mask in Latin American countries.

| Variables | Odds Ratio | Std. err. |  | Variables | Odds Ratio | Std. err. |  |
| --- | --- | --- | --- | --- | --- | --- | --- |
| **Sex (Men)** |  |  |  | **Political ideology (Right)** |  |  |  |
| Women | 1.483 | (0.086) | *** | Center-Right | 1.264 | (0.07) | * |
|  |  |  |  | Center-Left | 1.273 | (0.108) | ** |
| **Age (18-25)** |  |  |  | Left | 0.850 | (0.144) | * |
| 26-40 | 1.185 | (0.086) | * | None | 1.227 | (0.133) |  |
| 41-60 | 1.570 | (0.126) | *** | Missing | 1.094 | (0.127) |  |
| 61+ | 1.562 | (0.165) | *** |  |  |  |  |
| **Education (Incomplete Basic)** |  |  |  | **Country (Brazil)** |  |  |  |
| Basic | 1.119 | (0.112) |  | Argentina | 0.601 | (0.117) | * |
| Secondary | 1.428 | (0.15) | *** | Bolivia | 0.179 | (0.03) | *** |
| Tertiary | 2.161 | (0.262) | *** | Chile | 1.382 | (0.326) |  |
| **Labor activity (self-employed)** |  |  |  | Colombia | 0.501 | (0.092) | *** |
| Employee | 1.429 | (0.116) | *** | Costa Rica | 1.251 | (0.281) |  |
| Unemployed | 1.310 | (0.134) | * | Dominican Rep. | 0.503 | (0.092) | *** |
| Retired | 1.706 | (0.283) | ** | Ecuador | 0.892 | (0.182) |  |
| Inactive | 1.041 | (0.075) |  | El Salvador | 0.933 | (0.191) |  |
| Student | 1.381 | (0.209) | * | Guatemala | 0.976 | (0.197) |  |
| **Social Class (High)** |  |  |  | Honduras | 0.662 | (0.124) | * |
| Medium | 1.227 | (0.13) |  | México | 0.190 | (0.032) | *** |
| Low | 1.054 | (0.106) |  | Nicaragua | 0.341 | (0.06) | *** |
| Missing | 0.898 | (0.13) |  | Panamá | 0.649 | (0.125) | * |
| **Religion (Evangelical)** |  |  |  | Paraguay | 0.401 | (0.072) | *** |
| Catholic | 1.300 | (0.094) | *** | Perú | 0.659 | (0.125) | * |
| Other | 1.067 | (0.149) |  | Uruguay | 0.788 | (0.156) |  |
| None | 0.796 | (0.068) | ** | Venezuela | 2.070 | (0.534) | ** |
|  |  |  |  | **Constante** | 6.433 | (1.415) | *** |
|  |  |  |  | **N** | 19988 |  |  |

\*\*\* P<0.001; \*\* P<0.01, \* P<0.05

Regarding religion, Catholics have a higher chance of wearing the mask than Evangelicals (OR=1.300; p-value<0.001) while people who declare no religion use it to a lesser extent (OR=0.796; p-value <0.05). Results on political ideology indicate that people from the center-right (OR=1.264; p-value<0.05) and center-left (OR=1.273; p-value<0.01) use the mask to a greater extent than right-wing people. However, there are no significant differences between people who is on the right with respect to those on the left or without political ideology. Compared to Brazil, the reference category, the only country where the use of masks is significantly higher is Venezuela (OR=2.070; p-value<0.01), although Chile and Costa Rica also showed non-significant higher rates. In contrast, the use of the mask is lower in Bolivia, Colombia, the Dominican Republic, Honduras, Mexico, Nicaragua, Panama, Paraguay and Peru.

## Discussion

Here we draw a profile of people who commonly used a mask between October and November 2020, before the vaccination campaign started in Latin America. The results of this analysis are consistent with previous studies that indicate that men are more prone to risky behaviors than women (11). Although youngsters are less likely to use masks, both the middle-age and oldest-age groups showed similar predisposition, indicating that the age perception of the risk started earlier that the actual risk (12). Findings that having a higher educational level implies a greater use of masks are consistent with previous studies where higher educational levels are associated to healthier behaviors engagements (4).

Regarding occupational categories, the fact that retirees or pensioners have a greater use of the face mask than those who are self-employed is related to the fact that older people are the most vulnerable group to the virus (3). Additionally, students might be more likely to use them due to the mandatory use of face masks in educational centers. We also find that employees, as well as the unemployed, are more likely to wear a mask than those who are self-employed. In this sense, it should be noted that, although we control for the current occupational situation, the survey does not have variables that allow us to control which people can perform their daily lives without having to leave their home, where the use of a face mask would not be so necessary. Social class does not show significant results, but this might be a limitation of the self-perceive question to capture this dimension. Unlike countries like the US, where political orientation shapes prevention behaviors (5), the results of our study indicate that there is no left-right polarization of mask use in Latin America. Interestingly, centrist people are the most likely to wear a mask. In this sense, religion, which has also been present in discussions about the pandemic (13), also shapes prevention behaviors. Those belonging to the Evangelical religion wear a mask to a lesser extent than Catholics. These results open many questions about the role of religions in promoting health behaviors and the arguments that certain religions establish against medical discourse. Finally, Venezuela, Chile, Costa Rica and Brazil are the countries in Latin America where the use of a mask is highest. In the case of Brazil, a lack of rapid and equitable responses from the government might have led the population to increase their prevention attitudes against the virus (14). Regarding Venezuela, the country has deepened its political and economic crisis since COVID-19, and there has been a lack of credibility from the citizenship in the reported cases because of the shortage of virus tests (15).

The use of the face mask to prevent COVID-19 infection has been uneven across individual, regional, cultural and ideological factors in Latin America. The analysis showed the importance of understanding the demographic and socioeconomic profile of those individuals more prone to use preventive measures to stop the spread of the pandemic in the region, and the critical role of social forces–through religion and political ideas–in shaping health-related attitudes in a health emergency crisis.

## Data Availability

Data upon the research is done is freely available and ensures the confidentiality and privacy of the participants in accordance with relevant laws and regulations.

https://www.latinobarometro.org/latContents.jsp

